# Attitudes to phage therapy among Australian infectious diseases physicians

**DOI:** 10.1101/2023.07.03.23292153

**Authors:** Martin Plymoth, Stephanie A. Lynch, Ameneh Khatami, Holly A. Sinclair, Jessica C. Sacher, Jan Zheng, Ruby CY. Lin, Jonathan R. Iredell

## Abstract

Due to the rise in antimicrobial resistance (AMR), there has been an increased interest in phage therapy to treat multi-drug resistant infections. In Australia, phage therapy is predominantly used in small clinical studies or for compassionate use, however, despite its potential expansion in modern medicine, the perception of phage therapy among medical professionals remains largely unknown. Therefore, we conducted a national survey of Australian infectious diseases and clinical microbiology advanced trainees and specialists to assess their knowledge, areas of interest, and concerns around the use of phage therapy in clinical practice in Australia. Our survey received 92 responses from infectious diseases and clinical microbiology professionals across all states of Australia. The majority of those surveyed believed that the current national plan for controlling AMR is inadequate and that phage therapy may be an effective solution; with 97% of respondents indicating that they would consider using phage therapy meeting established guidelines for purity and safety (United States Food and Drug Administration and/or European Union guidelines). The respondents indicated a preference for bespoke therapy, with Gram-negative pathogens highlighted as priority targets. Alongside the phage therapy delivery protocols, therapeutic phage monitoring (TPM; like therapeutic drug monitoring (TDM)) was considered important. Cystic Fibrosis, lung-infections, prosthetic device related infections, and infections among patients following transplantation and/or immunosuppression were highly ranked in terms of priorities for clinical syndromes. Accessibility was highlighted as a barrier to phage therapy, specifically timely access (72%) and logistics of phage procurement and administration (70%). Altogether, these results suggest the support of phage therapy among infectious diseases and clinical microbiology advanced trainees and specialists in Australia, and highlights areas of focus and priority in order to advance phage therapy in modern medicine.

## Introduction

For over a century, phage therapy has been used to treat infectious diseases, although largely overshadowed by the discovery and development of antibiotics ^[1]^. With the rise in antimicrobial resistance (AMR), almost 5 million deaths globally were associated with resistant organisms in 2019 ^[2]^. It is predicted that by 2050, antibiotic-resistant infections will become the leading cause of death worldwide, resulting in approximately 10 million deaths annually ^[3]^. To address this, there has been increasing interest in the therapeutic application of phages, in particular for multi-drug resistant infections ^[1, 4]^.

Like other therapeutic agents, phage therapy will fall under regulation by the Australian Therapeutics Goods Administration (TGA), requiring adherence to strict manufacturing standards and demonstration of safety in clinical trials ^[5]^. Small clinical studies focusing on compassionate use of bacteriophages have been performed or are currently underway in Australia ^[5-7]^. With the potential for expansion of phage therapy in human medicine, perceptions towards this among relevant medical professionals remains largely unknown. A better understanding of this is necessary to inform future clinical studies and protocols.

Previous studies among the general public and certain patient populations have shown initial poor awareness, but high levels of support for, and acceptance of, phage therapy post briefing ^[8, 9]^. To the best of our knowledge, there have been no studies investigating knowledge, attitudes, and perceptions regarding phage therapy among infectious diseases clinicians, clinical microbiologists or trainees in such fields. Similarly, there have been no investigations regarding priorities for phage therapy implementation, or perceived barriers against this. In this cross-sectional study, we conducted a survey of Australian infectious diseases and clinical microbiology advanced trainees and specialists’ knowledge, areas of interest, and concerns around the use of phage therapy in clinical practice in Australia.

## Method

The study was based on a scoping qualitative survey attempting to identify priority areas for phage therapy which was conducted among infectious diseases and microbiology trainees and physicians between March 31^st^ and April 10^th^, 2021 (78 replies; **Supplement 1**). Based on replies to this survey, a quantitative survey containing 16-questions was developed with further input from experts in the field, including adult and paediatric infectious diseases physicians with established experience in phage therapy.

The survey was disseminated to infectious diseases physicians via the Australian Society of Infectious Diseases (ASID) OzBug mailing list (1,068 members, including non-prescribers). It was further advertised through local networks of infectious diseases clinicians. All participants were asked to agree to study participation with an electronic informed consent form. Survey results were collected via an online survey platform (JotForm Enterprises, San Francisco, CA) between 2 November 2021 and 30 October 2022. Appropriate statistical analysis was performed using IBM SPSS Statistics for Windows, version 27 (IBM Corp., Armonk, N.Y., USA), including Kruskal-Wallis H test, Mann-Whitney U test, and Wilcoxon matched-pairs signed-rank test. A p-value <0.05 was considered statistically significant.

Ethical approval was obtained from Western Sydney Local Health District Human Research Ethics Committee (2022/ETH00432). See **Supplement 1** for ethics application.

## Results

One hundred and six replies were collected during the survey period. Four identified as non-prescribers, and ten identified as trainees or specialists in fields other than infectious diseases or clinical microbiology and were excluded from further analysis. Among the remaining 92 respondents, 80 (87.0%) were specialists within the field of infectious diseases and/or microbiology, and a further 12 (13.0%) were advanced trainees. Participants were from across Australia, and approximately reflected the distribution of physicians across the country, although no respondents originated from the Australian Capital Territory (New South Wales 29%; Victoria 32%; South Australia 9%; Queensland 14%; Tasmania 1%; Western Australia 11%; Northern Territory 2%) ^[10]^.

Awareness of phage therapy as either adjunct or an alternative to antibiotics was scored on a Likert scale (1-5; 1 – minimal knowledge, 5 – expert knowledge) with most respondents admitting to a moderate level of knowledge of phage therapy (median 3/mode 3, interquartile range [IQR] 2-3; **figure 1**). No difference in self-reported awareness was identified based on participants’ geographic location (p=0.940); however, infectious diseases physicians in training positions reported lower awareness than specialists (median 2, IQR 1.25-3 vs. median 3, IQR 2-3.75, respectively; p=0.014).

**Figure 1.**
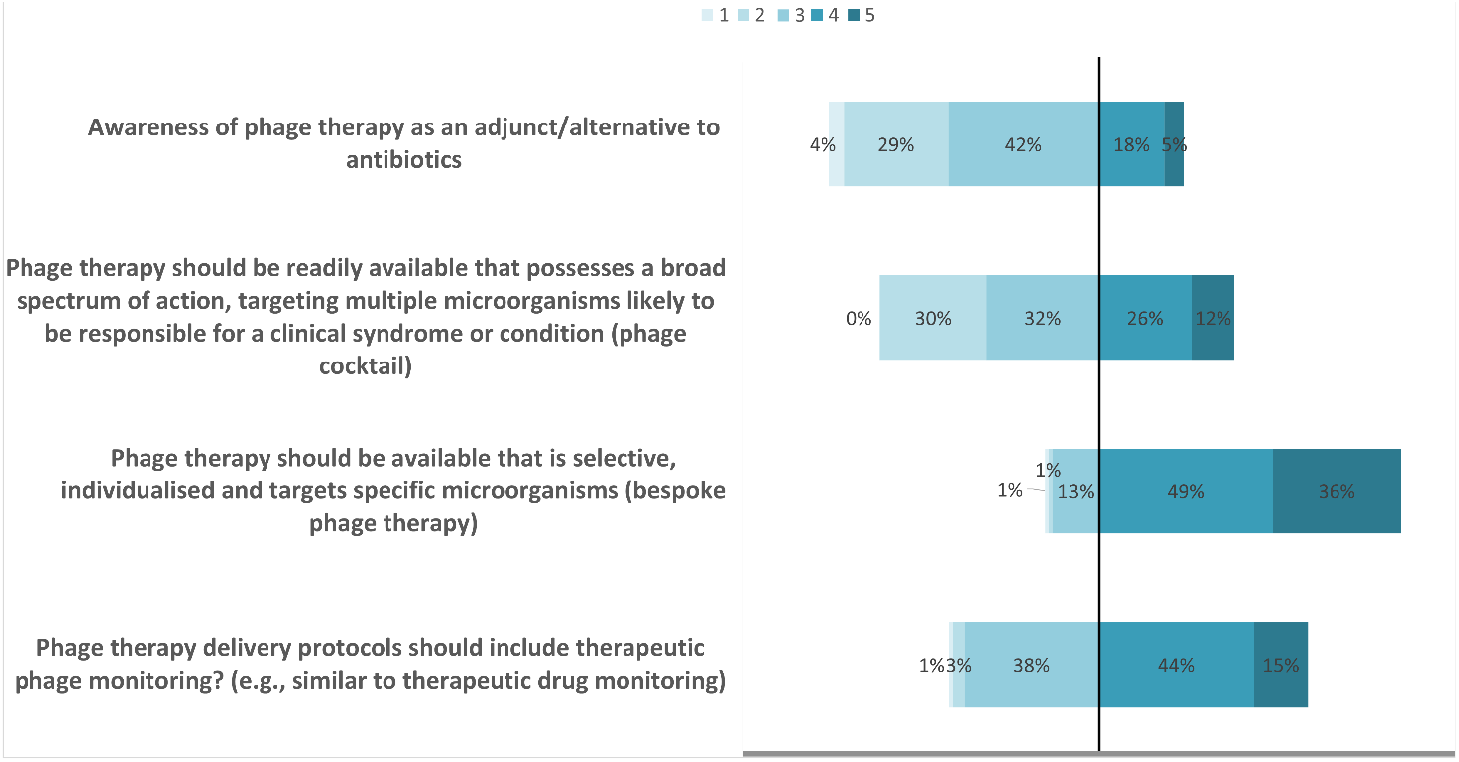
Knowledge, attitudes, and perceptions around phage therapy among Australian infectious diseases and microbiology clinicians (n=92). Ordinal scale: 1=lowest, 5=highest.

The majority of respondents (n=76, 83%) believed that Australia‘s national AMR action plan is currently inadequate in controlling the spread of AMR in Australia and around two-thirds (65%) believed phage therapy could be an effective solution to AMR. Most respondents (89; 97%) would consider using phage therapy meeting established guidelines for purity and safety (United States Food and Drug Administration and/or European Union guidelines); and the same number would participate in a clinical trial of phage therapy for their patients if given adequate support and resources. Most respondents had identified patients (multiple patients, n=35; 38%, or single patient n=28; 30%) who would have benefited from targeted phage therapy within the last year.

Perception of the relative importance of phage therapy availability as a combination ‘cocktail’ formulation or bespoke therapy was ranked on a Likert scale (1-5), with apparent preference for bespoke phage therapy (median 3, mode 2, IQR 2-4 for cocktails vs. median 4, mode 4, IQR 4-5 for bespoke; p<0.001). Therapeutic monitoring of phage therapy was also considered important by respondents (**figure 1**).

### Key areas for phage therapy development

Prioritised areas for targeted phage therapy (specific microorganisms and clinical syndromes) are displayed in **figure 2a**. Gram-negative organisms (*Pseudomonas aeruginosa*, Enterobacterales, *Acinetobacter baumannii*, and *Burkholderia* spp.) ranked highly as did mycobacterial species including *Mycobacterium abscessus*. Gram-positive organisms (*Enterococcus* spp. and *Staphylococcus aureus*) were least prioritised. Other non-listed target microorganisms identified by respondents included *Neisseria* spp. (n=2), *Sternotrophomonas* spp. (n=1), *Nocardia* spp. (n=1), *Brucella* spp. (n=1), *Coxiella* spp (Q fever) (n=1), *Achromobacter* spp. (n=1), *Clostridium difficile* (n=1), Coagulase-negative staphylococci (n=1), *Streptococcus pneumoniae* (n=1), *Helicobacter pylori* (n=1), and *Mycoplasma* spp. (n=1).

**Figure 2.**
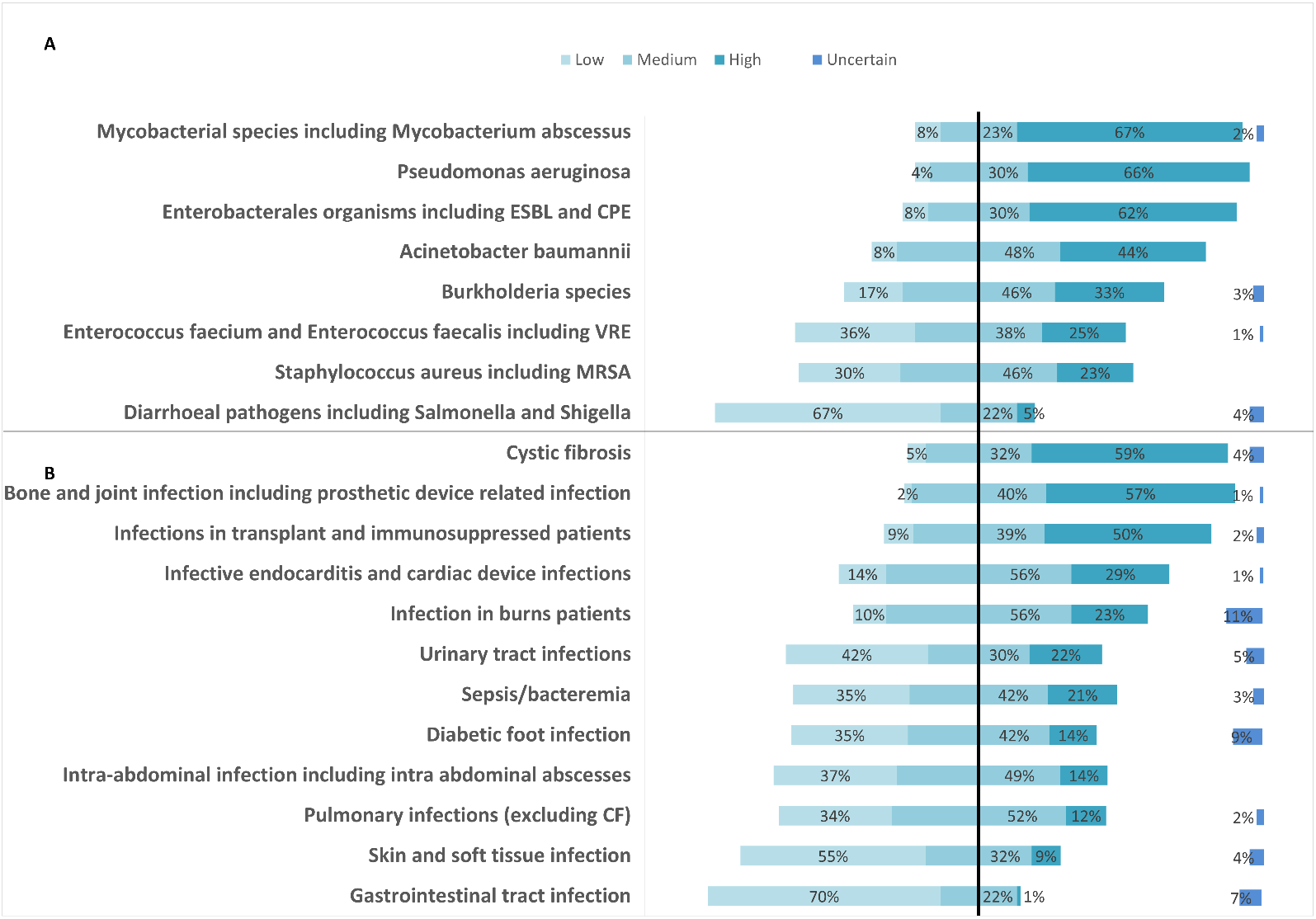
Prioritization areas for phage therapy development for (a) microorganisms; and (b) clinical syndromes by infectious diseases and microbiology clinicians (n=92); E. coli: *Escherichia coli*; ESBL: Extended-spectrum beta-lactamases; CPE: Carbapenemase-producing Enterobacterales; VRE: Vancomycin-resistant Enterococci; MRSA: Methicillin-resistant Staphylococcus aureus; CF: cystic fibrosis

Cystic fibrosis-related lung infections, bone and joint infections, prosthetic device related infections, and infections among patients following transplantation and/or immunosuppression were highly ranked in terms of priorities for clinical syndromes (**figure 2b**), while skin and gastrointestinal infections were least prioritized. Other clinical syndromes identified by respondents included vascular graft infections (n=4), and central nervous system infections (n=3). The selection of ‘uncertain’ to the order of priority was generally higher among the clinical syndromes listed than microorganisms listed.

Useful modes of delivery and administration of phage therapy were identified as follows: intravenous (85, 92%), inhalation (56, 61%), oral (56, 61%), instillation into body cavities and/or organs (32, 47%), phage-coated devices including prosthetic devices and indwelling catheters (36, 39%), and topical (31, 34%).

### Barriers to implementing phage therapy

Concerns regarding timely access to phage therapy, logistics of phage procurement and administration, as well as efficacy were primarily highlighted by respondents (72%, 70%, 58% of all respondents, respectively), while recruitment into clinical trials and bacterial resistance development were less concerning (23%, 20%, respectively; **Figure 3**).

**Figure 3.**
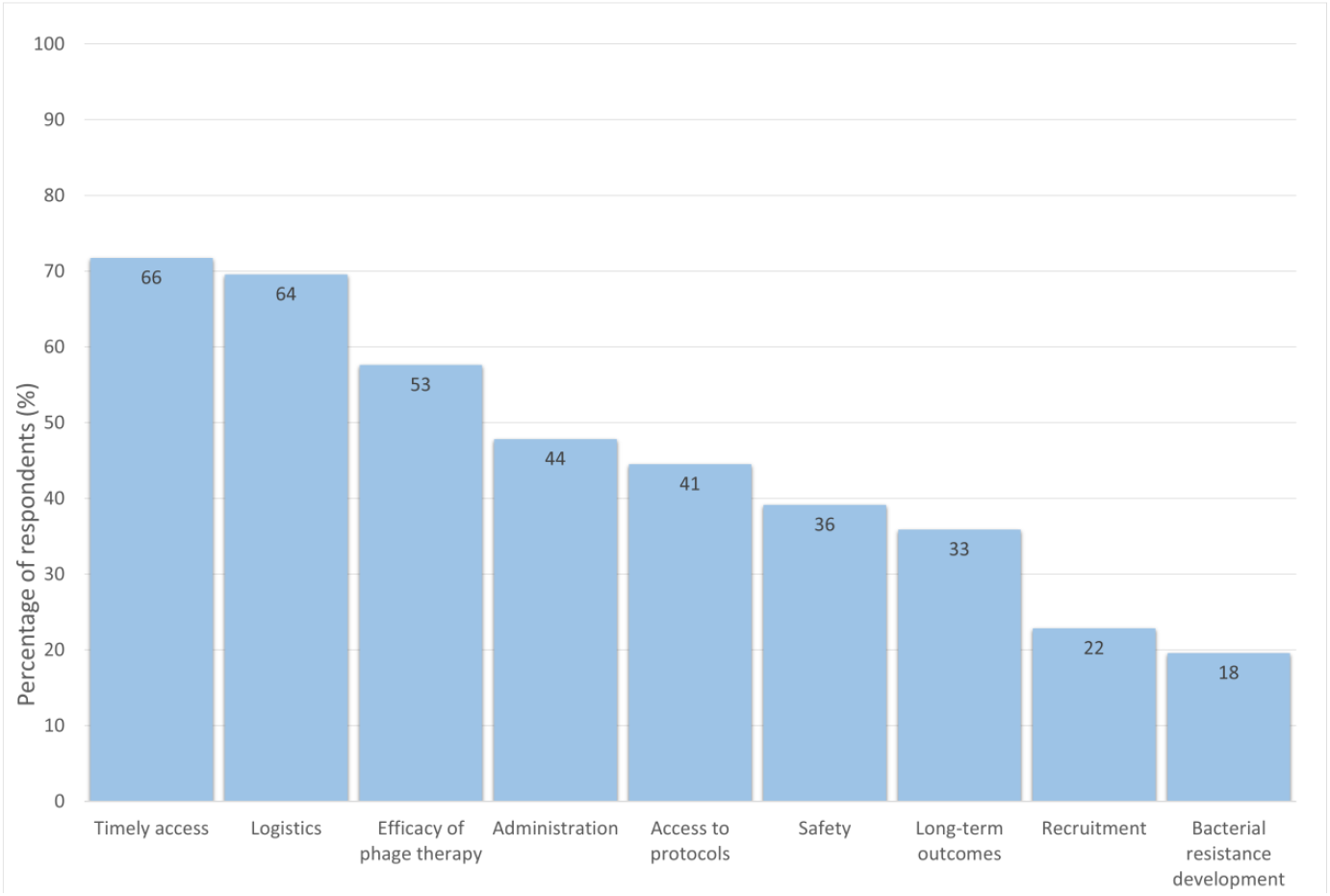
Primary considerations and perceived barriers against implementation of phage therapy, as identified by infectious diseases and microbiology clinicians. Absolute number in graph boxes.

## Discussion

In this study we report priorities for phage therapy in terms of microorganisms and clinical syndromes as identified by Australian infectious diseases and microbiology clinicians. In this study we further highlight the perceived concern by clinicians in current efforts to control the spread of AMR in Australia. This data also highlights their support for phage therapy as an adjunct or alternative to current antibiotics regimens and as a potential solution to this issue.

Prioritisation of phage therapy targeting Gram-negative organisms among respondents corresponds to the World Health Organisation (WHO) list of antibiotic-resistant bacteria that pose the greatest threat to human health, which identifies carbapenemase-producing *A. baumannii, P. aeruginosa*, and *Enterobacteriaceae* as critical priority pathogens for research and development ^[11]^.

A strong focus on Cystic Fibrosis, and its associated pathogens, as well as inhaled formulations of phage therapy was noted among many respondents. This aligns with the review of emerging pathogens such as *M. abscessus* that are identified as important targets for which antibiotic treatment options are limited ^[12]^. Furthermore, *P. aeruginosa* remains a poor prognostic factor in Cystic Fibrosis patients, especially with extensively drug-resistant strains ^[13]^. Introduction of effective new treatment regimens targeting such pathogens could lead to better outcomes for these individuals ^[14]^.

Interestingly, while bone and joint infections and prosthetic device-related infections were highly prioritised by respondents, targeted phage therapy towards *S. aureus*, the most frequent isolated pathogen in these infections, was less so. This could reflect the availability of conventional therapeutic options in these infections (antibiotics combined with surgical interventions such as irrigation, debridement and/or removal of infected prosthetic devices). However, where surgical interventions have failed, or are impractical, lifelong antibiotic suppressive therapy is often required. Case reports have shown successful outcomes using phages targeting *S. aureus* in prosthetic joint infections, suggesting they can potentially disrupt biofilm formation ^[15, 16]^ and provide additional benefit in selected cases.

Timely access to bacteriophages was identified as a critical barrier to implementation. This could be improved by a regulated framework supporting phage banks along with magistral preparations and personalised patient-specific therapies ^[17]^. Clinical collaboration, standardisation of protocols and development of national phage and pathogen biobanks, and a national surveillance program, as advocated by the Australian Phage Network are key ^[18]^. Despite concerns about timely access, clinicians indicated a preference for targeted ‘bespoke’ phage therapies for specific infections, as compared to broad-spectrum phage cocktails, highlighting the need for companion diagnostics to ensure effective phage therapy. This signifies the clinical need to continue to develop personalized phage matching, and production of phages targeted towards bacterial strains causing infection in a specific patient. Furthermore, development of standardized therapeutic monitoring for these therapies should be prioritized.

Limitations of this study include the relatively small sample size and uptake of the questionnaire, with only a minority of the estimated 600 infectious diseases physicians responding ^[19]^. The sample size lacked power for further sub-analysis. Furthermore, uncertainty in replies regarding prioritisation of some clinical syndromes (for example, infections in cystic fibrosis and burns patients) was higher than that regarding prioritisation of microorganisms, suggesting limited exposure to phage therapy and an understanding of its role in polymicrobial infections, as observed in other countries ^[20]^. With most Australian physician being hospital based, and phage therapy currently requiring close follow-up during its administration period, rural and remote medicine aspects were not fully considered in this study. Lastly, while limiting respondents to Australia gave a focused insight into the local clinical needs, it did not give a representation of Australasia as a whole, with other regions facing unique challenges in terms of logistics and access to medications.

In conclusion, our survey on phage therapy identified that physicians around Australia expressed support for properly conducted and supported clinical trials of phage therapy. Priority research areas identified suggest an unmet need in antibiotic resistant and prosthetic device infections.

## Data Availability

All data produced in the present study are available upon reasonable request to the authors

## Author contributions

Survey concept: JI, RL

Questionnaire design: JI, RL, MP, SL, AK, HS, and JS, JZ HREC/methods design: SL, RL

Data analysis: SL and MP

Manuscript writing: SL, MP, RL

Members of the Phage Australia network were consulted.

**Supplement 1**

