## Supplement 1 for "Attitudes to phage therapy among Australian infectious diseases physicians"

Martin Plymoth[1,2], Ruby C Y Lin[2,3,4], David L Paterson[5,6], Jonathan R Iredell[1,2,3*]

1. Westmead Hospital, Sydney, New South Wales, Australia
2. School of Medicine, The University of Sydney, Sydney, New South Wales, Australia
3. Centre for Infectious Diseases and Microbiology, Westmead Institute for Medical Research, Sydney, New South Wales, Australia
4. School of Medical Sciences, University of New South Wales, Sydney, New South Wales, Australia
5. Centre for Clinical Research, The University of Queensland, Brisbane, Queensland, Australia
6. Royal Brisbane and Women's Hospital, Brisbane, Queensland, Australia

**Main text** 562 words

For over a century, phage therapy has been identified as a method to treat infectious diseases, although largely overshadowed by the discovery and development of antibiotics. With emergence of antimicrobial resistance, research on therapeutic bacteriophages has once again become of interest [1]. In Australia, phage therapy is regulated by the Australian Therapeutics Goods Administration (TGA), requiring adherence to strict manufacturing standards and demonstration of safety in clinical trials [2]. While small clinical studies and compassionate use of bacteriophages have been performed in Australia, the perception among relevant medical professionals towards phage therapy remains largely unknown and a better understanding necessary is to inform future clinical studies and protocols [2, 3].

In this cross-sectional study we conducted a semi-quantitative survey aimed towards Australian infectious diseases advanced trainees and specialists using the Australasian Society for Infectious Diseases (ASID) mailing list, which had 962 Australian members (including clinical microbiologists, scientist, infection control practitioners, public health physicians, sexual health physicians, veterinarians) at time of posting the questionnaire on March 31st, 2021.

We received 78 replies between March 31st and April 10th 2021. Most (76; 97%) of respondents identified as Australian infectious diseases physicians or trainees, the majority (72; 95%) working either solely or partially in the public sector. Correspondence roughly reflected the distribution of physicians across the country*.

Awareness of phage therapy as an adjunct/alternative to antibiotics was scored on a Likert scale (0-10), with a mean of 5.0 (95% CI 4.5-5.5). Most (60; 79%) of respondents believed that phage therapy meeting American Food and Drug Administration (FDA) guidelines for purity would be safe to administer to patients, while 17 (22%) were unsure [4]. The vast majority (69; 91%) would consider using phage therapy if available and 72 (95%) supported recruitment into clinical trials with prepared phage therapy, given adequate support.

Primary concerns to address were identified from keywords in short free text answers (**Fig 1a**); these included timely access to appropriate phages (16; 21%), uncertainty about efficacy (11; 15%) and safety (10; 13%), clinical trial protocols (9; 12%), administration and dosing (7; 9%), and resistance development (6; 8%). Concerns about biosecurity, effect on microbiome, coadministration with antibiotics, and spectrum of broad-range bacteriophage cocktails were also described. Similarly, the highest priority research areas (**Fig 1b**) were identified as being Gram-negative microorganisms (17; 22%) and prosthetic joint and device infections (15; 20%), with Pseudo*monas aeruginosa* (10; 13%), Non-Tuberculous Mycobacteria (NTM)/*Mycobacterium abscessus* (10; 13%), and *Staphylococcus aureus* (6; 8%) particularly mentioned. Other pathogens (*Burkholderia* spp., *Acinetobacter* *baumannii*, *Helicobacter pylori, Mycobacterium tuberculosis*) and diseases (cystic fibrosis, bone and joint infections, urinary tract infections, burns, diabetic foot infections, endocarditis) were further identified as areas of interest for bacteriophage therapy.

In conclusion, our survey in phage therapy identified interested physicians around Australia who expressed support for properly conducted and supported clinical trials. Priority research areas identified suggest unmet need in antibiotic resistant infections and prosthetic device infections.

Timely access to bacteriophages could be improved by a regularly framework supporting phage banks along with magistral preparations and personalised patient-specific therapies [4]. This could be facilitated by clinical collaboration, standardisation of protocols and development of national phage and  pathogen biobanks and a national surveillance program, as advocated by the Australian Phage Network [5].

---

***** New South Wales 26 (34%); Victoria 17 (22%). Queensland 12 (16%); West Australia 8 (11%); South Australia 7 (9%); Northern Territory 4 (5 %); and 1 each from Tasmania and ACT (1%)

**Ethics**

Our survey among non-dependent adult medical professionals was deemed low-risk and unlikely to cause offense. Data was collected anonymously. As per recommendation from the Human Research Ethics Manual, no ethics approval was requested from the Human Research Ethics Committee (HREC) of Australia. The survey was conducted in accordance with professional ethics, including the Australian Code of Responsible Conduct of Research and The WMA Declaration of Helsinki.

**Contributors**

All authors contributed to the conceptualization and methodology of the study. MP and JRI designed the survey. MP collected and analysed responses. MP, RCYL and JRI wrote the original draft. DLP reviewed the manuscript. All authors approved the final manuscript.

**Declaration of interest**

RCYL, DLP, JRI are members of the Australian Phage Network. DLP has received research grants from MSD, Pfizer and Shionogi and is on the advisory boards of Spero, MSD, Pfizer, QPex, Entasis, Janssen, GSK and Accelerate. We declare no further conflicts of interest.

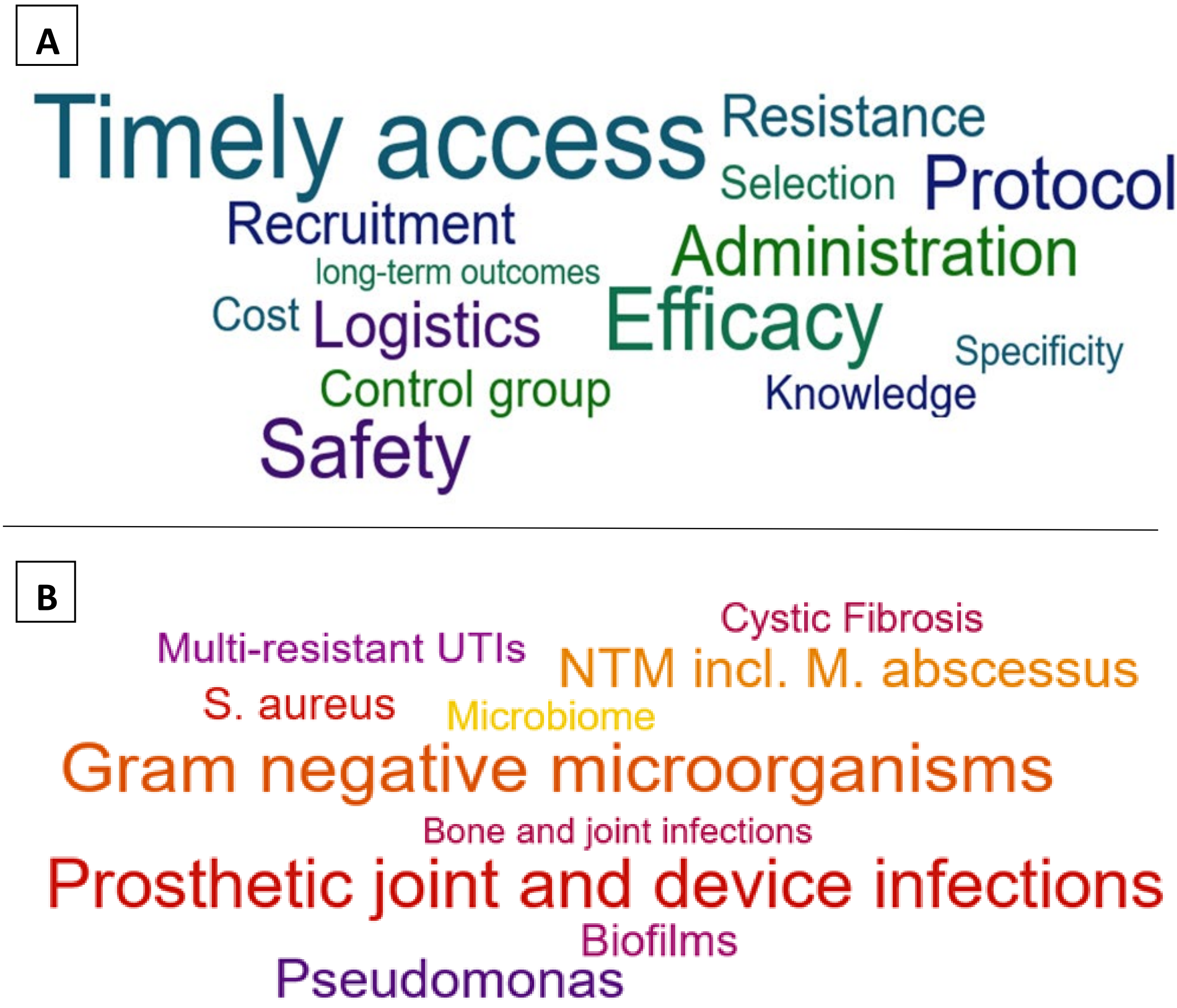

**Fig 1.** Highest priority (**A**) concerns with regards to clinical trials; and (**B**) infections/pathogens research areas for phage therapy, as perceived by Australian infectious diseases physicians.

Total respondents: n=76. NTB: Non-Tuberculous Mycobacteria; M. abscessus: *Mycobacterium abscessus*; S. aureus: *Staphylococcus aureus*; UTIs: Urinary tract Infections

#### Human Research Ethics Application

**Application Management Information**
**Application ID:** 2022/ETH00432
**Created date:** 13/04/2022
**Originating Application ID:** 2022/ETH00432
**This is the earliest application from which this application (2022/ETH00432) was copied.*
**Parent Application ID:** 2022/ETH00432
**This is the immediate predecessor from which this application (2022/ETH00432) was copied.*
**Version Number:** 2
**Application submitted to:** Western Sydney Local Health District; Western Sydney Local Health District Human Research Ethics Committee.

**The applicant has requested that this ethics application be considered under the Greater than low risk review pathway.**

### Section 1 – Core Information

#### Pre-application conditions

**The applicant/s have acknowledged that:**

1. The HREA has been designed for ethics review of human research, as defined in the National Statement.
2. Adequate resources must be available to conduct this research project.
3. All relevant institutional polices pertaining to the conduct of this research project should be considered and adhered to.
4. Research activities must not commence until ethics approval (and site authorisation, if appropriate) has been provided.

#### Project Overview

**Q1.1 Project Title:**

Perception of phage therapy among Australian authorised prescribers

**Q1.2 Summary of the research project:**

(Bacterio)phages are bacterial viruses that are recognised as a potential adjunct or alternative treatment to antibiotics, against highly drug-resistant infections in humans. Whilst there has been an increase in the use of such therapy in human medicine, uncertainties remain within the medical field. Therefore, we intend to distribute an anonymous survey to authorised prescribers including Infectious Disease (ID) clinicians, clinical microbiologists and trainees within the field, to understand their attitudes towards phage therapy. No identifying information will be captured from the respondents and there is a small risk of duplicate entries. The survey intends to identify the major concerns surrounding phage therapy and the priorities for phage therapy among authorised prescribers. The anonymous survey responses will be analysed to create a report that will inform Phage Australia and the medical professional community of the clinical demand for phage therapy and the formulations that will best suit clinical practice.

**Q1.3 Which category/ies of research best describes the project?**

Clinical Sciences - 1103

**Q1.4 In what environments will the research be conducted?**

| |
|---|
| Research institute(s) |

**Q1.5 What organisation/entity has overall responsibility for this project?**

| |
|---|
| Sponsor type: Institution/Investigator-initiated<br>Sponsor name: Western Sydney Local Health District<br>Westmead Institute for Medical Research |

**Q1.6 Describe how this research project is currently, or will be, funded.**

| |
|---|
| No funding required. Investigator initiated. |

**Q1.7 Anticipated starting date of the research project:**

| |
|---|
| As soon as ethics and any other relevant approvals have been provided. |

**Q1.8 Anticipated duration of the research project:**

| |
|---|
| 6 Months |

#### Project Team

*Name:* **Prof. Jonathan Iredell**

**Q1.9.4 Email Address:**

**Q1.9.5 Is this person the contact person for this application?**

No

**Q1.9.6 Is this person a student on this project?**

No

**Q1.9.7 Institutional affiliation and position:**

Director of Centre for Infectious Disease and Microbiology, Westmead Institute for Medical Research/Senior Infectious Diseases & Microbiology Practitioner, Westmead Hospital

**Q1.9.8 Staff ID** *(optional)***:**

**Q1.9.9 ORCID Identifier** *(optional)***:**

**Q1.9.10 Position on the research project:**

Co-ordinating Principal Investigator/Researcher

**Q1.9.12 Research activities Prof. Jonathan Iredell will be responsible for:**

Principle investigator of project. Survey question design and implementation. Survey distribution to clinical networks.

**Q1.9.13 Expertise relevant to the research activity:**

Experienced infectious diseases specialist and clinical microbiologist with experience in clinical phage therapy in Australia including treatment of adults. Past president of AMR Australia, Senior infectious diseases staff at Westmead Hospital (former head of department), Director of Phage Australia and Centre for Infectious Diseases and Microbiology

*Name:* **Dr. Stephanie Lynch**

**Q1.9.4 Email Address:**

**Q1.9.5 Is this person the contact person for this application?**

Yes

| | |
|---|---|
| **Q1.9.5.1 Email Address:** | |

| | |
|---|---|
| **Q1.9.5.2 Telephone Number:** | 0423204152 |
| **Q1.9.5.3 Mailing Address** | 2/22 Robert Street, Telopea, NSW, 2117 |

**Q1.9.6 Is this person a student on this project?**

No

**Q1.9.7 Institutional affiliation and position:**

Westmead Institute for Medical Research, Postdoctoral Researcher

**Q1.9.8 Staff ID** *(optional)***:**

**Q1.9.9 ORCID Identifier** *(optional)***:**

**Q1.9.10 Position on the research project:**

Investigator/Researcher

**Q1.9.12 Research activities Dr. Stephanie  Lynch will be responsible for:**

Question design and survey creation. Data collection and analysis. Stakeholder engagement.

**Q1.9.13 Expertise relevant to the research activity:**

Experienced in science communication, survey designs and engagement with key stakeholders, knowledge in phage biology

*Name:* **Assoc. Prof. Ruby Lin**

**Q1.9.4 Email Address:**

**Q1.9.5 Is this person the contact person for this application?**

No

**Q1.9.6 Is this person a student on this project?**

No

**Q1.9.7 Institutional affiliation and position:**

Westmead Institute for Medical Research, Business Development Lead
Deputy Director, Phage Australia

**Q1.9.8 Staff ID** *(optional)***:**

**Q1.9.9 ORCID Identifier** *(optional)*:

**Q1.9.10 Position on the research project:**

Associate/Assistant/Sub-/Co- Investigator/Researcher

**Q1.9.12 Research activities Assoc. Prof. Ruby Lin will be responsible for:**

Survey design, data analysis, Participant recruitment/survey distribution through clinical networks, engagement with stakeholders, facilitate Phage Australia clinical and biologics networks

**Q1.9.13 Expertise relevant to the research activity:**

Deputy Director of Phage Australia, Genomic Chair of Scientific Advisory Group at Westmead Research Precinct (3 medical research institutes and 2 public hospitals), She managed and is scientific lead for first in human clinical trial using GMP-grade Staph aureus phage cocktail in adults.

*Name:* **Dr. Jessica Sacher**

**Q1.9.4 Email Address:**

**Q1.9.5 Is this person the contact person for this application?**

No

**Q1.9.6 Is this person a student on this project?**

No

**Q1.9.7 Institutional affiliation and position:**

Postdoctoral Researcher, Westmead Institute for Medical Research.

**Q1.9.8 Staff ID** *(optional)*:

**Q1.9.9 ORCID Identifier** *(optional)*:

**Q1.9.10 Position on the research project:**

Investigator/Researcher

**Q1.9.12 Research activities Dr. Jessica Sacher will be responsible for:**

Survey design. Data collection. Engagement with key stakeholders.

**Q1.9.13 Expertise relevant to the research activity:**

co-founder of Phage Directory, experience with scientific communication, survey designs, data collection, phage biology and engagement with stakeholders

*Name:* **Dr. Martin Plymoth**

**Q1.9.4 Email Address:**

**Q1.9.5 Is this person the contact person for this application?**

No

**Q1.9.6 Is this person a student on this project?**

No

**Q1.9.7 Institutional affiliation and position:**

Western Sydney Local Health District, infectious diseases clinical fellow

**Q1.9.8 Staff ID** *(optional)***:**

**Q1.9.9 ORCID Identifier** *(optional)***:**

**Q1.9.10 Position on the research project:**

Investigator/Researcher

**Q1.9.12 Research activities Dr. Martin Plymoth will be responsible for:**

infectious diseases, survey design, clinical cohort, data collection and data analysis

**Q1.9.13 Expertise relevant to the research activity:**

experience in clinical infectious diseases, clinical microbiology, conducted first prescribers survey on phage therapy

*Name:* **Mr. Jan Zheng**

**Q1.9.4 Email Address:**

**Q1.9.5 Is this person the contact person for this application?**

No

**Q1.9.6 Is this person a student on this project?**

No

**Q1.9.7 Institutional affiliation and position:**

Westmead Institute for Medical Research, Bioinformatician

**Q1.9.8 Staff ID** *(optional)***:**

**Q1.9.9 ORCID Identifier** *(optional)***:**

**Q1.9.10 Position on the research project:**

Investigator/Researcher

**Q1.9.12 Research activities Mr. Jan Zheng will be responsible for:**

Survey design, data collection, data analysis, engagement with key stakeholders, channels of communication

**Q1.9.13 Expertise relevant to the research activity:**

co-founder of Phage Directory, experience with scientific communication, survey designs, data collection, phage biology and engagement with stakeholders

*Name:* **Dr. Holly Sinclair**

**Q1.9.4 Email Address:**

**Q1.9.5 Is this person the contact person for this application?**

No

**Q1.9.6 Is this person a student on this project?**

No

**Q1.9.7 Institutional affiliation and position:**

Queensland local health district, infectious diseases fellow

**Q1.9.8 Staff ID** *(optional)***:**

**Q1.9.9 ORCID Identifier** *(optional)***:**

**Q1.9.10 Position on the research project:**

Investigator/Researcher

**Q1.9.12 Research activities Dr. Holly Sinclair will be responsible for:**

survey design, data collection and data analysis, engage with stakeholders

**Q1.9.13 Expertise relevant to the research activity:**

infectious diseases, clinical microbiology

*Name:* **Dr. Ameneh Khatami**

**Q1.9.4 Email Address:**

**Q1.9.5 Is this person the contact person for this application?**

No

**Q1.9.6 Is this person a student on this project?**

No

**Q1.9.7 Institutional affiliation and position:**

senior staff at Chidren's Hospital Westmead senior lecturer at University of Sydney deputy directory of Phage Australia

**Q1.9.8 Staff ID** *(optional)***:**

**Q1.9.9 ORCID Identifier** *(optional)***:**

**Q1.9.10 Position on the research project:**

Investigator/Researcher

**Q1.9.12 Research activities Dr. Ameneh Khatami will be responsible for:**

Survey design, data collection and analysis, engagement with stakeholders

**Q1.9.13 Expertise relevant to the research activity:**

Infectious disease staff at CHW (paediatrics), had experience treating kids with phage

*Name:* **Prof. Jonathan Iredell**

**Q1.9.4 Email Address:**

**Q1.9.5 Is this person the contact person for this application?**

No

**Q1.9.6 Is this person a student on this project?**

No

**Q1.9.7 Institutional affiliation and position:**

Director of Centre for Infectious Disease and Microbiology, Westmead Institute for Medical Research/Senior Infectious Diseases & Microbiology Practitioner, Westmead Hospital

**Q1.9.8 Staff ID** *(optional)*:

**Q1.9.9 ORCID Identifier** *(optional)*:

**Q1.9.10 Position on the research project:**

Chief Investigator/Researcher

**Q1.9.12 Research activities Prof. Jonathan Iredell will be responsible for:**

Principle investigator of project. Survey question design and implementation. Survey distribution to clinical networks.

**Q1.9.13 Expertise relevant to the research activity:**

Experienced infectious diseases specialist and clinical microbiologist with experience in clinical phage therapy in Australia including treatment of adults. Past president of AMR Australia, Senior infectious diseases staff at Westmead Hospital (former head of department), Director of Phage Australia and Centre for Infectious Diseases and Microbiology

---

#### Disclosure of interests

**Q1.10 Do any members of the research team (including persons not listed in this application), have any financial or non-financial interests related to this research?**

No

---

#### Restrictions

**Q1.11 Are there any restrictions or limits on publication of data or dissemination of research outcomes of this project?**

No

---

#### Evaluations

**Q1.12 Has the scientific or academic merit of the research project been evaluated?**

No

**Q1.13 Has this research project had prior ethics review?**

No

**Q1.14 Will any further or additional specialised review of this application be sought?**

No

---

#### Setting of research

**Q1.15 Will this project be conducted at multiple sites?**

No

**Q1.16 Will separate institutional approvals or authorisations be required prior to commencing research at each site?**

No

---

### Section 2 – Research Details and Participants

**Q1.17 The following research methods will be used in the research project:**

| Research Method | Status |
|---|---|
| Action research | ✗ |
| Biospecimen analysis research | ✗ |
| Data linkage research | ✗ |
| Ethnographic research | ✗ |
| Epidemiological research | ✗ |
| Interventional/Clinical Trials research | ✗ |
| Observational research | ✗ |
| Survey/Interview/Focus Group research | ✓ |
| Textual analysis research | ✗ |
| None of the above | ✗ |

**Q1.18 The research will be conducted with the following:**

| Participation | Status |
|---|---|
| Human beings (via active participation), including their associated biospecimens and/or data. | ✓ |
| Human biospecimens only | ✗ |
| Data associated with human beings only (i.e. as the primary object of research) | ✗ |

**Q1.19 The research will involve the following participants:**

| Participants | Status |
|---|---|
| Women who are pregnant and the human fetus | ✗ |
| Children and young people | ✗ |
| People highly dependent on medical care who may be unable to give consent | ✗ |
| People with a cognitive impairment, intellectual disability or mental illness | ✗ |
| People in dependent or unequal relationships | ✗ |
| People who may be involved in illegal activities | ✗ |
| People in other countries | ✗ |
| Aboriginal and Torres Strait Islander peoples | ✗ |

#### Method Specific Questions

---

##### Survey/Interview/Focus Group Research

**M8.1 What process/es will your research project use?**

Surveys

**M8.2 How will you engage with your participants?**

|Indirectly, via an online provider

**M8.3 How will personal identifiers be retained or removed over the course of your project?**

The survey being distributed to Australian prescribers is entirely anonymous as there is no identifying information collected and the responses cannot be traced back to the respondent. Therefore, no idenfiers will need to be retained or removed over the course of this project.

**M8.4 Will participants have the opportunity to review or edit their responses or contributions prior to data analysis or publication?**

Yes

**M8.4.1.1 Indicate the relevant section/s of your Project Description that detail this opportunity.**

This information is documented under >Methods >Data collection “Participants will be able to review their responses prior to submission and subsequent data analysis by scrolling through all questions and altering responses prior to clicking the submit button.”

**M8.5 Is it foreseeable that your project will explore topics that may cause distress for participants?**

No

#### Participant Specific Questions

#### Recruitment Questions

**Q2.1.1 Indicate how you will identify and recruit participants for your research, referencing any relevant sections of your Project Description/Protocol as appropriate.**

As stated in >Methods >Survey distribution and participation; "The intended audience of this survey are clinicians in Australia with Australian Health Practitioner Regulation Agency (APHRA) registration, working at major tertiary teaching hospitals, therefore, the email will be sent to ID clinicians, clinical microbiologists and trainees across Australia by:  Specific clinicians within our network that will distribute the email containing the survey link to their professional network (known as snowballing).  The PI of this study will ask key clinical organisations across Australia to distribute the email containing survey links to clinical members on their email lists (e.g., OzBug through Australasian Infectious Diseases Society (https://www.asid.net.au/members/ozbug-2) a defined infectious diseases clinical group.  A public link of the survey will be shared on social media platforms; Twitter and LinkedIn." Participants will recieve all required documentation via email. Participation is voluntary and the participant can withdraw at anytime. The link will remain open for 6 months from HREA approval, and the participant can choose when they fill out the survey in their own time.

**Q2.1.2 How will your recruitment strategy take account of the ethical considerations relevant to the specific people you are recruiting?**

According to pg16 of Element 2 of National Statement Chapter 3.1 participants give consent by ticking the consent button at the begining of the survey and returning response to this survey. This survey is designed to be anonymous and data cannot be reverse engineered to identify the participant. As such, there is no recruitment strategy except for an e-mail notification that such survey exists within the Australian infectious diseases professional groups.

---

#### Consent Questions

**Q2.2.1 Indicate by reference the relevant section/s of your Project Description/Protocol that address/es consent.**

As stated in >Methods >Survey design; “The survey will be accompanied by a Patient Information Consent Forms (PICFs) (see attachment), where participants will be required to tick ‘yes’ to the pre-survey consent question to give their consent to participate. If participants select ‘no’ to the pre-survey consent question, the survey responses will be invalid." All PICFs and relevant informaiton will be distributed to the participant via email. The survey will remain open for 6 months, therefore, participants can fill out the survey in their own time if they choose to participate. The survey requires the participant to check a box to recieve participant consent and will be stored in the survey database.

**Q2.2.2 Will you be obtaining consent from some or all participants to participate in the research?**

Yes for all participants

**Q2.2.2.1 What is the scope of consent that you will be seeking?**

Specific

**Q2.2.2.2 How will consent be obtained?**

Written

Implied

**Q2.2.2.3 Are you proposing to obtain consent using limited disclosure?**

No

**Q2.2.3 Are family members, authorised representatives or any others involved in the participants' decision to participate in the research?**

No

**Q2.2.4 Will there be an opportunity to confirm or re-negotiate consent during the research project?**

Yes

**Q2.2.6 Describe any ethical considerations related to the approach to consent that you will be seeking and your strategies for addressing and managing these issues.**

The survey will be distributed to clinicians in Australia with Australian Health Practitioner Regulation Agency (APHRA) registration, working at major tertiary teaching hospitals, therefore, the email will be sent to ID clinicians, clinical microbiologists and trainees across Australia. Therefore, the questions will be easily understood by participants. The participant can revoke or re-negotiate consent at anytime throughout the survey, by choosing not to fill out the survey and closing their browser, this way, no answers will be saved.

**Q2.2.7 Are you proposing to use an opt-out approach with respect to some or all of the participants?**

No

**Q2.2.8 Are you requesting a waiver of the requirement for consent with respect to some or all participants?**

No

#### Risk Questions

**Q 2.3.1 Describe the risks and burdens associated with your research, referencing any relevant sections of your Project Description as appropriate.**

There is no risk or burdens associated with this survey to participants, as the questions aim to understand what authorised prescribers think about phage products, phage therapy (health services) and the entities (companies/institutes) who will provide the phages, and these topics should not cause any distress to the participant.

**Q 2.3.2 Describe how these risks will be mitigated and managed.**

This survey is a low risk survey, risks have been mitigated by designing questions that should not cause risk or harm or distress to the participants.

#### Benefit Questions

**Q2.4.1 Describe the benefits associated with your research, referencing any relevant sections of your Project Description as appropriate.**

The outcome will benefit the medical community on the following: 1. Perception of phage therapy amongst prescribers in Australia in the first instance in this AMR global crisis (we've only conducted a smaller scale survey back in March 2021). 2. The clinical indications given by the participants (clinical community) will inform Phage Australia regarding the type of phages needed at the bedside (bespoke or large volume). This will then inform production of phage products. 3. This information will also inform curation of different reference set of bacteria panel required to screen appropriate phages toward therapy. 4. The information captured will advance research in phage biology, specifically in research for matching phages, synergy testing with antibiotics, phage and pathogen - all of this can inform clinical decisions for difficult to treat infections. 5. This information is useful in terms of pandemic preparedness/surveillance towards biobanking and stockpiling phages and pathogen collection for future use.

**Q2.4.2 Explain how benefits of this research justify any risks or burdens associated with the research.**

This is an anonymous survey, there is low to no risk associated but the benefits outweigh any risk or burden associated with this research outcome.

**Q2.4.3 How will you manage participants' expectations of the perceived benefit of participating in the research?**

We have facilitated a clinical network i.e., a group of clinicians and researchers who are interested in using phage therapy as an antimicrobial resistant solution in a clinical setting. The outcome of this survey will inform the participants and would meet their expectations of all the benefits outlined above in Q2.4.1. Specifically, clinicians at different hospitals would observe a specific common infection, and results from the survey can inform local manufacturing of bespoke phages to meet this demand. In the case of an outbreak then large scale GMP production of phages that is centralised at Westmead would meet the demand. In addition to this, we have also facilitated a biobanking network which will meet the expectations of the participants that specific phages are accessible to meet patient demand.

### Section 3 – Data and Privacy

#### Data Characteristics

**Q3.1 Indicate the type of information/data you will be collecting for this project.**

Not personal information

**Q3.2 Indicate the type of information/data you will be using in this project:**

Not personal information

**Q3.3 Indicate the degree of identifiability of information/data you will be collecting for this project.**

Non-identifiable information

**Q3.4 Indicate the degree of identifiability of information/data you will be using in this project.**

Non-identifiable information

**Q3.5 Describe any ethical considerations relating to the collection and/or use of the information/data in this project.**

We have a data management plan, risk mitigation plan as well as a feasibility plan under the Phage Australia framework. This survey falls under this framework. Collection of this data is anonymous and we have a plan in place to ensure security of the data collected. This data will not be shared to a third party.

**Q3.6 Identify the source/s of the information/data that you will be collecting and/or using in this project.**

Individual participants and/or relatives or associates of participants

**Q3.7 Describe any ethical considerations relating to the source of information/data as indicated in the response to the previous question.**

No data collected will be identifiable to the individuals who participated in this survey. No data will be collected that can identify medical/health/mental health records of the participants. This is an anonymous survey that participant can opt out at any time during this survey - incomplete survey will not be recorded. The questions outlined in the survey relate to normal clinical practice by the participants.

**Q3.8 Was the information/data that you are using previously collected for a purpose other than research?**

No

---

#### Activities Planned for/with Data

**Q3.9 Do you plan to disclose any personal information/data in this project to a third party?**

No

**Q3.10 How will you protect the privacy of participants and non-participants in any notes and/or publications arising from your research?**

No personal data will be collected. The survey form is designed to be anonymous. All form data will be stored on dedicated servers, encrypted by 256-bit SSL, PCI DSS Level I and HIPAA certification. Raw data will be retained for 5 years post-publication. All data collected is unidentifiable, with no questions asking for identifiable data

**Q3.11 Are there any restrictions on your ability to assure the confidentiality of participants?**

No

**Q3.12 Do you plan to share any individual research results obtained during this research to the participants?**

No

**Q3.13 Describe how you will handle any secondary or incidental findings that arise from the analysis of personal information/data.**

The survey is designed with a specific outcome with mostly binary answers, yes or no thus incidental correlation is minimal i.e., there will be no link to clinically actionable issues because no personal data is collected. No genomics, imaging and non-medical data will be collected.

**Q3.14 Describe how the information/data will be stored, accessed, archived and/or destroyed.**

All form data will be stored on dedicated servers, encrypted by 256-bit SSL, PCI DSS Level I and HIPAA certification. Raw data will be retained for 5 years post-publication. All data collected is unidentifiable, with no questions asking for identifiable data

**Q3.15 Describe any ethical considerations relating to the storage of, access to or destruction of information/data in this project.**

The survey aims to collect quantitative data which will be automatically collated on the JotForm Tables function, with the required data fields set as columns (see data collection form attached). Survey responses collected in JotForm Tables database are password protected, by which, only investigators of this study can access. All form data will be stored on dedicated servers, encrypted by 256-bit SSL, PCI DSS Level I and HIPAA certification. Raw data will be retained for 5 years post-publication. All data collected is unidentifiable, with no questions asking for identifiable data.

**Q3.16 Will the outcomes of this project be disseminated to the participants?**

Yes

**Q3.16.1.1 Describe how the outcomes of the project will be disseminated to the participants, or refer to the relevant section/s of your Project Description/Protocol which deals with this matter.**

This survey aims to understand the attitudes of authorised prescribers towards phage therapy, as well as determine the clinical needs of phage therapy within the health care system across Australia. Thus the results of this survey will be disseminated to the participants. Currently, there is no published data that explores the in depth perception of phage therapy among this cohort of health professionals, therefore, such research findings would be a novel area of work in the phage therapy field. By disseminating results from this survey, the intention is to inform research groups across Australia on the priorities and receptiveness of phage therapy from ID clinicians, clinical microbiologists and trainees to allow for more efficient research and guide subsequent research efforts.

**Q3.16.1.2 Describe any ethical considerations relating to any dissemination of outcomes to the participants.**

The survey is anonymous and no personal identifiable data will be collected. The results of the survey may change the perception of prescribers towards phage therapy and pathogen surveillance management - such information contributes to better clinical management thus we do not foresee any ethical issues.

**Q3.17 Describe any foreseeable future activities for which information/data collected and/or used in this project may be made available.**

We intend to disseminate the results of this survey to the wider community other than the prescribers. This will increase discussion on using phage as a therapeutic option in the clinics in Australia. We also foresee increased discussion amongst clinical trainees and researchers to conduct more efficient research and increase research effort into phage therapy. In addition, we foresee that results of this survey can inform the Therapeutics Goods Administration (TGA) of the clinical community’s perception towards phage therapy, Currently phage therapy is legislated under TGA's special access scheme.

**Q3.18 Describe any ethical considerations relating to the planned or possible future use of information/data in this project.**

This is a anonymous survey that will reflect the perception of prescribers' attitude towards phage therapy. We do not foresee any potential obligations to act on the findings in the future. There is no IP ownership associated with this survey. No personal data is collected so participant privacy is preserved at all times.

### Section 4 – Attachments and Declarations

#### Attachments

The following documents have been attached to this HREA.

##### Project Description/Protocol

| |
|---|
| See attachment *07032022 HREA project description.docx* |

##### Other attachments

| **Type** | **Attachment File Name** | **Attachment Description** |
|---|---|---|
| Questionnaire | *20042022_Prescriber survey questionaire.pdf* | Updated survey questions |
| Participant Consent Form | *[Tracked] 20042022_WSLHD_PICF_Version_1.2.doc* | Prescriber survey PICF Version 1.2 Tracked |
| Letter of Invitation | *04022022_Prescriber_survey_distribution_email.docx* | Survey distribution email |
| Other, Please specify | *04022022_Perception_of_phage_therapy_data_collection_form.xlsx* | Survey collection form |
| Ethics application (HREA or other) | *04022022_HREA_Submission_Checklist_Version_1.docx* | HREA submission checklist |
| Other | *11032022_Proof_of_fee_payment.pdf* | Proof of submission fee payment |
| Participant Consent Form | *20042022_WSLHD_PICF_Version_1.2.doc* | Prescriber survey PICF Version 1.2 Clean |
| Other, Please specify | *26042022_SAC Review Responses_RL_JI.docx* | SAC HREC response letter |
| | Project Registration | The output from the |

#### Investigator Team Declarations

**The research team has certified that:**

- All information in this application and supporting documentation is correct and as complete as possible;
- I have read and addressed in this application the requirements of the National Statement and any other relevant guidelines;
- I have familiarised myself with, considered and addressed in this application any relevant legislation, regulations, research guidelines and organisational policies;
- All relevant financial and non-financial interests of the project team have been disclosed; and
- In the capacity of a supervisor, as applicable, I have reviewed this application and I will provide appropriate supervision to the student(s) in accordance with the arrangements specified in this application and those associated with the student's educational program.

| |
| --- |
| ☑ Certified |

**Title:** Perception of phage therapy among Australian authorised prescribers
**Version Number:** Version 1

#### Project Team Roles & Responsibilities:

- Prof. Jon Iredell, Westmead Hospital, Senior Infectious Diseases & Microbiology Practitioner
    - Principal investigator/project lead. Survey design, Participant recruitment/survey distribution through clinical networks. Director of Phage Australia.
- Assoc. Prof. Ruby Lin, Westmead Institute for Medical Research, Business Development Lead.
    - Survey design, data analysis, Participant recruitment/survey distribution through clinical networks, engagement with stakeholders, facilitate Phage Australia clinical and biologics networks.
- Dr. Stephanie Lynch, Westmead Institute for Medical Research, Postdoctoral Researcher.
    - Survey design and creation. Data collection and analysis. Engage with stakeholders
- Dr. Jessica Sacher, Westmead Institute for Medical Research, Postdoctoral Researcher.
    - Survey design. Data collection. Engagement with key stakeholders.
- Mr. Jan Zheng, Westmead Institute for Medical Research, Bioinformatician.
    - Survey design, data collection, data analysis, engagement with key stakeholders, channels of communication
- Dr. Martin Plymoth, Western Sydney Local Health District, infectious diseases clinical fellow
    - Infectious diseases, survey design, clinical cohort, data collection and data analysis
- Dr. Holly Sinclair, Queensland local health district, infectious diseases fellow
    - survey design, data collection and data analysis, engage with stakeholders
- Dr. Ameneh Khatami, senior staff at Chidren's Hospital Westmead senior lecturer at University of Sydney deputy directory of Phage Australia
    - Survey design, data collection and analysis, engagement with stakeholders

#### Resources:

- Project is investigator initiated and funded. No external resources or funding required.

#### Background:

Modern medicine has adopted two approaches to control the myriad of infectious diseases, vaccination for prevention and antimicrobials for treatment of such diseases [1]. However, many of the currently used antimicrobials are no longer effective in treating infections due to antimicrobial resistance (AMR). AMR is the evolution of microorganisms, particularly bacteria, to survive applications of antimicrobial compounds, including antibiotics [2]. With the global increase in AMR, statistics predict that by 2050, antibiotic-resistant infections will become the leading cause of death worldwide, resulting in approximately 10 million deaths annually [3]. Therefore, the World Health Organisation (WHO) has identified a list of antibiotic-resistant bacteria that pose the greatest threat to human health, many of which significantly burden the healthcare system and are 'priority pathogens' for new antimicrobial strategies [4]. While antibiotics are one such antimicrobial strategy to treat bacterial infections, the development of antibiotic classes with novel modes of action is declining, therefore, there is a need for alternative therapeutics [1]. Phage therapy offers a third approach to infectious disease control. Phage therapy is the therapeutic application of (bacterio)phages, which are viruses that prey on bacteria [5]. One such advantage of phages is that they are effective against antibiotic-resistant bacterial strains, offering a last defence against otherwise untreatable infections [5]. In addition, phage therapy provides numerous advantages within a clinical setting. For example, unlike antibiotics, each phage is highly precise in the specific bacteria that it targets, meaning that treatment has fewer effects on the healthy bacteria in the human body, thus less likely to cause antimicrobial selective pressure and secondary infections [ref]. Importantly, pure phage preparations have shown to be non-toxic to humans and such phage preparations can be used on their own or in combination with other phages, therefore, improving the safety and efficacy of phage therapy in human medicine [6–8]. With increased attention towards phage therapy, there are now many studies that support the safety and efficacy of phage therapy in human medicine, typically in the form of compassionate use, which is the use of phages outside of clinical trials whereby all other therapeutic options have been exhausted [9]. With the significant expansion of phage therapy in human medicine, it is essential to understand the clinical needs and the attitudes of clinicians towards phage therapy, to better inform the research and progression of phage therapy. To the best of our knowledge, there is currently no literature investigating the perceptions of phage therapy from infection disease (ID) clinicians, clinical microbiologists or trainees in such fields, or the clinical barriers and priorities of phage therapy. Interestingly, there are a few studies that have explored the patients experience and perceptions of phage therapy, of which, the data extracted from the survey could inform healthcare professionals and policy makers of the patient acceptability of phage therapy [10,11]. Such data from Australian prescribers would be invaluable in determining the progress of phage therapy in Australian modern medicine. Therefore, this study aims to use a quantitative survey to help interpret authorised prescribers' perceptions about phage therapy in Australia.

**Aim:** To understand the attitudes towards phage therapy and the clinical needs of phage therapy among authorised prescribers across Australia.

#### Project Design:

##### *Survey design*

An anonymous online market research survey was developed to understand what authorised prescribers think about phage products, phage therapy (health services) and the entities (companies/institutes) who will provide the phages. The choice of an online survey compared to hard copy surveys allows quick access to a large sample size without the constraints on time and/or location. Additionally, the respondent data is easy to access, process and analyse. The survey questions were designed as close-ended questions toward a quantitative research framework, to ensure that responses could be statistically analysed or used to infer patterns, trends or correlations between data points. The content of the survey questions was carefully phrased by a group of internal clinicians who have previously worked on phage therapy research and have successfully treated patients with phage therapy, therefore, increasing the validity and reliability of the data collected. The survey was developed using a sub-set of 16 carefully selected questions and were input into JotForm Enterprises (San Francisco, CA) (https://form.jotform.com/212990934520053). The 16 quantitative questions contained a selection of tick-boxes, rating scales, ranking scales and free-text responses, which is estimated to take 3 minutes to fill out. The survey will be accompanied by a Patient Information Consent Forms (PICFs) (see attachment), where participants will be required to tick 'yes' to the pre-survey consent question to give their consent to participate. If participants select 'no' to the pre-survey consent question, the survey responses will be invalid. The survey format and questions have been reviewed by phage experts within the field, along with clinicians across Australia, prior to its dissemination.

##### *Survey distribution and participation*

A digital format of the survey (https://form.jotform.com/212990934520053) will be distributed to the intended audience via email (see attachment) with the PICFs (see attachment) attached to the email. The intended audience of this survey are clinicians in Australia with Australian Health Practitioner Regulation Agency (APHRA) registration, working at major tertiary teaching hospitals, therefore, the email will be sent to ID clinicians, clinical microbiologists and trainees across Australia by:

- Specific clinicians within our network that will distribute the email containing the survey link to their professional network (known as snowballing).
- The PI of this study will ask key clinical organisations across Australia to distribute the email containing survey links to clinical members on their email lists (e.g., OzBug through Australasian Infectious Diseases Society

(https://www.asid.net.au/members/ozbug-2) a defined infectious diseases clinical group.

- A public link of the survey will be shared on social media platforms; Twitter and LinkedIn.

The survey will remain open for responses for 6 months, with the aim of collecting responses from >150 prescribers. Reminder emails and updates will be sent at regular intervals to optimise response rate among intended participants. Investigators within the research group have previously conducted a similar survey from Feb-March 2021, to a similar population. Therefore, this prior experience gives us confidence in achieving a large enough sample size for data analysis. To note, this cohort of infectious diseases clinicians are currently dealing with COVID response thus response rate maybe biased.

***Data collection***

The survey aims to collect quantitative data which will be automatically collated on the JotForm Tables function, with the required data fields set as columns (see data collection form attached). Survey responses collected in JotForm Tables database are password protected, by which, only investigators of this study can access. All form data will be stored on dedicated servers, encrypted by 256-bit SSL, PCI DSS Level I and HIPAA certification. Raw data will be retained for 5 years post-publication. All data collected is unidentifiable, with no questions asking for identifiable data. Participants will be able to review their responses prior to submission and subsequent data analysis by scrolling through all questions and altering responses prior to clicking the submit button.

***Statistical analysis***

Survey responses collated in JotForm Tables will be downloaded into a password protected CSV file, opened in Microsoft Excel. Two investigators will be assigned to manually sort through the responses, and delete any invalid responses, whereby the respondent selected 'no' for the pre-survey consent question, or if the survey was submitted incomplete. Survey responses remaining after exclusion will undergo statistical analysis using SPSS as follows: Raw data will first be filtered by the state that the respondent is located and analysed to give a crude sense of the need for phage therapy in each state (based on question 7, 11 and 12). Raw data will be cross-tabulated to compare the answers to specific questions across the rest of the survey and responses will be analysed using a joint-distribution between two or more discrete variables (e.g. priority of pathogens to target and priority of clinical syndromes to treat). Statistical analysis for multivariate comparison (e.g. to infer correlation patterns) will involve the use of one-way ANOVA with Benjamini-Hochberg FDR post hoc adjustment. In all statistical analysis used, a P-value <0.05 would be considered significant. A summary of all analysed data will be presented using appropriate visualisations on Prism, GraphPad software. Such visualisation will include response percentages, response counts, pie charts and bar graphs.

#### Results, Outcomes and Future Plans:

In conclusion, our survey aims to understand the attitudes of authorised prescribers towards phage therapy, as well as determine the clinical needs of phage therapy within the health care system across Australia. Currently, there is no published data that explores the in-depth perception of phage therapy among this cohort of health professionals, therefore, such research findings would be a novel area of work in the phage therapy field. Data from the survey specifically aims to inform research groups across Australia on the priorities and receptiveness of phage therapy from ID clinicians, clinical microbiologists and trainees to allow for more efficient research and guide subsequent research efforts. Additionally, survey responses aim to inform the Therapeutics Goods Administration (TGA) of the clinical community's perception towards phage therapy, to further discussions surrounding the regulation of phage therapy in modern medicine.

### Participant Information Sheet/Consent Form

| | |
|---|---|
| **Title** | Perception of phage therapy among Australian authorised prescribers |
| **Coordinating Principal Investigator** | Prof Jonathan Iredell |
| **Site Principal Investigator** | Prof Jonathan Iredell |
| **Location** | |

#### What does my participation involve?

##### 1 Introduction

You are invited to take part in this clinical research survey. The research aims to understand the attitudes of authorised prescribers (Infectious disease clinicians, clinical microbiologists and trainees of the field) and the clinical needs of phage therapy.

This research is being conducted by Prof. Jon Iredell, Assoc. Prof. Ruby Lin, Dr. Stephanie Lynch, Dr. Jessica Sacher, Dr. Holly Sinclair, Dr. Martin Plymoth, Jan Zheng.

**Participation in this research is voluntary.** If you don't wish to take part, you don't have to, you may close the survey at any time and responses will not be recorded.
If you decide you want to take part in the research project, you will be asked to tick 'yes' to the pre-survey consent question within the survey. By ticking 'yes' it you are telling us that you:
• Understand what you have read
• Consent to take part in the research survey

**Responses are unidentifiable.** Questions through this survey do not ask for identifiable data and by participating in the survey, we will not be able to identify you.

**There are two qualifications to participate in this study.** (1) You are an authorised prescriber in Australia; (2) You select 'yes' to the pre-survey consent question.

**Please read this information carefully.** If you have any questions about the study, please contact Stephanie Lynch via.

##### 2 What is the purpose of this research?

| |
|---|
| Due to the rise in antimicrobial resistance (AMR) recent statistics predict that by 2050, antibiotic-resistant infections will become the leading cause of death worldwide, resulting in approximately 10 million deaths annually 3. To address this concern, there has been a recent increase in interest surrounding phage therapy, the therapeutic application of phages. With the significant expansion of phage therapy in human medicine, it is essential to understand the clinical needs and the attitudes of clinicians towards phage therapy, to better inform the research and progression of phage therapy. To the best of our knowledge, there is currently no literature investigating the perceptions of phage therapy from infection disease (ID) clinicians, clinical microbiologists or trainees in such fields, or the clinical barriers and priorities of phage therapy. Therefore, this study aims to use a quantitative anonymous questionnaire to survey clinicians in Australia, to understand the attitudes of authorised prescribers towards phage therapy, as well as determine the clinical needs of phage therapy within the health care system across Australia |

If you have any complaints about any aspect of the project, the way it is being conducted or any questions about being a research participant in general, then you may contact:

**Contact person**

| | |
|---|---|
| Name | *[Name]* |
| Position | *[Position]* |
| Telephone | *[Daytime Phone number] and [After Hours Phone number]* |
| Email | *[Email address]* |

**Reviewing HREC approving this research and HREC Executive Officer details**

| | |
|---|---|
| Reviewing HREC name | Western Sydney Local Health District (WSLHD) |
| HREC Executive Officer | Kellie Hansen |
| Telephone | (02) 8890 9007 |
| Email | |

**Complaints contact person**

| | |
|---|---|
| Position | *[Position]* |
| Telephone | *[Daytime Phone number] and [After Hours Phone number]* |
| Email | *[Email address]* |

**Site RESEARCH GOVERNANCE Office contact**

| | |
|---|---|
| Position | *[Position]* |
| Telephone | *[Phone number]* |
| Email | *[Email address]* |

### Consent Form - Adult providing own consent

| | |
|---|---|
| **Title** | Perception of phage therapy among Australian authorised prescribers |
| **Coordinating Principal Investigator** | Prof Jonathan Iredell |
| **Site Principal Investigator** | Prof Jonathan Iredell |
| **Location** | |

**<u>Declaration by Participant</u>**

I have read the Participant Information Sheet or someone has read it to me in a language that I understand.

I have had an opportunity to ask questions and I am satisfied with the answers I have received.

I freely agree to participate in this research project as described and understand that I am free to withdraw by closing the survey at any time.

By ticking ‘yes’ in the pre-survey consent question within the survey, I agree to the above declaration. See example of the pre-survey consent question below:

Pre-survey consent: I agree that I have read the informed consent document and agree to participate in this study. If you do not agree, please close this survey.

- ◯ Yes
- ◯ No

Phage Australia

### Perception of phage therapy among Australian authorised prescribers

This survey is intended for those that will prescribe phage therapy, including ID physicians, clinical microbiologists and trainees in the field. The reports and outcomes of this survey will aim to inform Phage Australia of the clinical demand for phage therapy and the formulations that will best suit clinical practice. Thank you for taking part in the survey, it should take approximately 3 minutes to complete. For more information, please head to https://phageaustralia.org/

Pre-survey consent: I agree that I have read the informed consent document and agree to participate in this study. If you do not agree, please close this survey.

- ◯ Yes
- ◯ No

Q1. What is your primary area of work/expertise? *

- ◯ Infectious diseases/clinical microbiology specialist
- ◯ Infectious diseases/clinical microbiology trainee
- ◯ Non-prescriber
- ◯ Other specialty/trainee

Now create your own Jotform - It's free! Create your own Jotform

- New South Wales
- Victoria
- South Australia
- Tasmania
- Queensland
- Northern Territories
- ACT
- West Australia

Q3. How well informed are you about phage therapy as an adjunct/alternative to antibiotics? *

1 2 3 4 5

I have no knowledge — I have expert/specialist knowledge

Q4. Would you consider using phage formulations that meet FDA and/or EU guidelines for purity and safety? *

- Yes
- No

Q5. Would you participate in a clinical trial on phage therapy for your patient, given adequate support/resourcing? *

- Yes
- No

Q6. What are your major concerns/doubts when it comes to clinical trials using phage therapy? (Check all that apply) *

Now create your own Jotform - It's free! Create your own Jotform

- [ ] Access to protocols
- [ ] Administration
- [ ] Bacterial resistance development
- [ ] Efficacy of phage therapy
- [ ] Logistics
- [ ] Long-term outcomes
- [ ] Recruitment
- [ ] Safety
- [ ] Timely access
- [ ] Other

Q7. Please rank the following organisms based on their priority for phage therapy? *

| | High priority | Medium priority | Low priority |
|---|---|---|---|
| *Enterobacterales (E. coli, Klebsiella spp., Enterobacter spp.) including ESBL and CPE.* | ○ | ○ | ○ |
| *Acinetobacter baumannii* | ○ | ○ | ○ |
| *Enterococcus faecium* and *Enterococcus faecalis* including VRE | ○ | ○ | ○ |
| *Staphylococcus aureus* including MRSA | ○ | ○ | ○ |
| Mycobacterial species including *Mycobacterium abscessus* | ○ | ○ | ○ |
| Diarrhoeal pathogens including *Salmonella* and *Shigella* | ○ | ○ | ○ |
| *Burkholderia species* | ○ | ○ | ○ |
| *Pseudomonas aeruginosa* | ○ | ○ | ○ |

Q8. If there is an organism you consider high priority that wasn't listed in Q7, please list it here

Q9. Phage therapy should be available that is selective, individualised and

Now create your own Jotform - It's free! Create your own Jotform

1 2 3 4 5

Strongly disagree Strongly agree

Q10. Phage therapy should be readily available that possesses a broad spectrum of action, targeting multiple microorganisms likely to be responsible for a clinical syndrome or condition (phage cocktail). *

1 2 3 4 5

Strongly disagree Strongly agree

Q11. Please select which clinical syndromes would benefit the most from phage therapy? *

| | High priority | Medium priority | Low priority |
|---|---|---|---|
| Bone and joint infection including prosthetic device related infection | ○ | ○ | ○ |
| Cystic fibrosis | ○ | ○ | ○ |
| Diabetic foot infection | ○ | ○ | ○ |
| Gastrointestinal tract infection | ○ | ○ | ○ |
| Infection in burns patients | ○ | ○ | ○ |
| Infections in transplant and immunosuppressed patients | ○ | ○ | ○ |
| Infective endocarditis and cardiac device infections | ○ | ○ | ○ |
| Intra-abdominal infection including intra abdominal abscesses | ○ | ○ | ○ |
| Pulmonary infection (excluding CF) | ○ | ○ | ○ |
| Sepsis/bacteremia | ○ | ○ | ○ |
| Skin and soft tissue infection | ○ | ○ | ○ |
| Urinary tract infections | ○ | ○ | ○ |

Q12. In the past year, have you come across a patient who you think may have

Now create your own Jotform - It's free! Create your own Jotform

- Yes, multiple patients
- Yes, a single patient
- No
- I am unsure

Q13. Which administration routes would you find useful for administering phage therapy? (Please select all that apply) *

- Inhalation
- Instillation (e.g., into organs or cavity)
- Intravenous
- Oral
- Phage-coated devices (including prostheses and indwelling catheters)
- Topically
- Other

Q14. Do you think phage therapy delivery protocols should include therapeutic phage monitoring? (e.g., Similar to therapeutic drug monitoring) *

1 2 3 4 5

Strongly disagree Strongly agree

Q15. Do you think Australia's antimicrobial resistance (AMR) strategy including the current antimicrobial stewardship programs are sufficient to prevent further AMR development? *

- Yes
- No

Q16. Do you believe phage therapy is a possible solution to prevent further AMR development? *

- Yes
- No

Submit

Now create your own Jotform - It's free! Create your own Jotform

Now create your own Jotform - It's free!

Create your own Jotform

| Submission Date | Pre-survey consent: I agree that I have read the informed consent document and agree to participate in this study. If you do not agree, please close this survey. | Q1. What is your primary area of work/expertise? |
|---|---|---|

| Q2. Which state/territory is your primary region of practice? | Q3. How well informed are you about phage therapy as an adjunct/alternative to antibiotics? | Q4. Would you consider using phage formulations that meet FDA and/or EU guidelines for purity and safety? |
|---|---|---|

| Q5. Would you participate in a clinical trial on phage therapy for your patient, given adequate support/resourcing? | Q6. What are your major concerns/doubts when it comes to clinical trials using phage therapy? (Check all that apply) | Q7. Please rank the following organisms based on their priority for phage therapy? >> Staphylococcus aureus including MRSA |
|---|---|---|

| Q7. Please rank the following organisms based on their priority for phage therapy? >> Enterococcus faecium and Enterococcus faecalis including VRE | Q7. Please rank the following organisms based on their priority for phage therapy? >> Enterobacterales (E. coli, Klebsiella spp., Enterobacter spp.) including ESBL and CPE. | Q7. Please rank the following organisms based on their priority for phage therapy? >> Pseudomonas aeruginosa |
|---|---|---|

| Q7. Please rank the following organisms based on their priority for phage therapy? >> Mycobacterial species including Mycobacterium abscessus | Q7. Please rank the following organisms based on their priority for phage therapy? >> Acinetobacter baumannii | Q7. Please rank the following organisms based on their priority for phage therapy? >> Burkholderia species |
| --- | --- | --- |

| Q7. Please rank the following organisms based on their priority for phage therapy? >> Diarrhoeal pathogens including Salmonella and Shigella | Q8. If there is an organism you consider high priority that wasn't listed in Q7, please list it here | Q9. Phage therapy should be available that is selective, individualised and targets specific microorganisms (bespoke phage therapy). |
|---|---|---|

| Q10. Phage therapy should be readily available that possesses a broad spectrum of action, targeting multiple microorganisms likely to be responsible for a clinical syndrome or condition (phage cocktail). | Q4A. Would you consider using phage formulations if it was produced as a certified GMP product? | Q11. Please select which clinical syndromes would benefit the most from phage therapy? >> Bone and joint infection including prosthetic device related infection |
|---|---|---|

| Q11. Please select which clinical syndromes would benefit the most from phage therapy? >> Cystic fibrosis | Q11. Please select which clinical syndromes would benefit the most from phage therapy? >> Diabetic foot infection | Q11. Please select which clinical syndromes would benefit the most from phage therapy? >> Gastrointestinal tract infection |
|---|---|---|

| Q11. Please select which clinical syndromes would benefit the most from phage therapy? >> Infection in burns patients | Q11. Please select which clinical syndromes would benefit the most from phage therapy? >> Infections in transplant and immunosuppressed patients | Q11. Please select which clinical syndromes would benefit the most from phage therapy? >> Infective endocarditis and cardiac device infections |
|---|---|---|

| Q11. Please select which clinical syndromes would benefit the most from phage therapy? >> Intra-abdominal infection including intra abdominal abscesses | Q11. Please select which clinical syndromes would benefit the most from phage therapy? >> Pulmonary infection (excluding CF) | Q11. Please select which clinical syndromes would benefit the most from phage therapy? >> Sepsis/bacteremia |
|---|---|---|

| Q11. Please select which clinical syndromes would benefit the most from phage therapy? >> Skin and soft tissue infection | Q11. Please select which clinical syndromes would benefit the most from phage therapy? >> Urinary tract infections | Q12. In the past year, have you come across a patient who you think may have benefited from phage therapy? |
|---|---|---|

| Q13. Which administration routes would you find useful for administering phage therapy? (Please select all that apply) | Q14. Do you think phage therapy delivery protocols should include therapeutic phage monitoring? (e.g., Similar to therapeutic drug monitoring) | Q15. Do you think Australia's antimicrobial resistance (AMR) strategy including the current antimicrobial stewardship programs are sufficient to prevent further AMR development? |
|---|---|---|

**Q16. Do you believe phage therapy is a possible solution to prevent further AMR development?**

**From:**
**Subject:** Phage Australia: 'Perception of phage th...

No images? Click here

Phage Australia

Dear colleagues,

We invite you, as an infectious disease physician and/or clinical microbiologist or trainee in the field, to participate in this **3 minute survey** on 'Perception of phage therapy among Australian prescribers'.

Your anonymous responses to this survey will inform **Phage Australia** of the **clinical demand for phage therapy** and the **formulations to best suit clinical practice**, for **more efficient phage therapy production**.

We encourage respondents to fill in the survey now, and **we will provide public access to the results within 6 months.**

**Please participate in our 'Perception of phage therapy among Australian prescribers' survey**

Please click this link to fill out the survey:
https://form.jotform.com/phage_therapy_prescriber

#### If you have any questions, please email us

If the link above doesn't work, please copy & paste this link into your preferred browser: https://form.jotform.com/212990934520053

Phage Australia

### Perception of phage therapy among Australian authorised prescribers

This survey is intended for those that will prescribe phage therapy, including ID physicians, clinical microbiologists and trainees in the field. The reports and outcomes of this survey will aim to inform Phage Australia of the clinical demand for phage therapy and the formulations that will best suit clinical practice. Thank you for taking part in the survey, it should take approximately 5 minutes to complete. For more information, please head to https://phageaustralia.org/

Pre-survey consent: I agree that I have read the informed consent document and agree to participate in this study. If you do not agree, please close this survey.

- Yes
- No

Q1. What is your primary area of work/expertise? *

- Infectious diseases/clinical microbiology specialist
- Infectious diseases/clinical microbiology trainee
- Non-prescriber
- Other specialty/trainee

Q2. Which state/territory is your primary region of practice? *

- Victoria
- South Australia
- New South Wales
- ACT
- West Australia
- Tasmania
- Queensland
- Northern Territories

Q3. In your opinion, how well informed are you about phage therapy as an adjunct/alternative to antibiotics? *

1 2 3 4 5

I have no knowledge — I have expert/specialist knowledge

Q4. Would you consider using phage formulations that meet FDA and/or EU guidelines for purity and safety? *

- Yes
- No

Q5. Would you participate in a clinical trial on phage therapy for your patient, given adequate support/resourcing? *

- ◯ Yes
- ◯ No

Q6. What are your major concerns/doubts when it comes to clinical trials using phage therapy? (Check all that apply) *

- ☐ Access to protocols
- ☐ Administration
- ☐ Bacterial resistance development
- ☐ Efficacy of phage therapy
- ☐ Logistics
- ☐ Long-term outcomes
- ☐ Recruitment
- ☐ Safety
- ☐ Timely access
- ☐ Other

Q7. In your opinion, how would you rank the following organisms based on their priority for phage therapy? *

| | High priority | Medium priority | Low priority | Uncertain |
|---|---|---|---|---|
| *Staphylococcus aureus* including MRSA | ◯ | ◯ | ◯ | ◯ |
| *Enterococcus faecium* and *Enterococcus faecalis* including VRE | ◯ | ◯ | ◯ | ◯ |
| *Enterobacterales (E. coli, Klebsiella spp., Enterobacter spp.) including ESBL and CPE.* | ◯ | ◯ | ◯ | ◯ |
| *Pseudomonas aeruginosa* | ◯ | ◯ | ◯ | ◯ |
| Mycobacterial species including *Mycobacterium abscessus* | ◯ | ◯ | ◯ | ◯ |
| *Acinetobacter baumannii* | ◯ | ◯ | ◯ | ◯ |
| *Burkholderia species* | ◯ | ◯ | ◯ | ◯ |
| Diarrhoeal pathogens including *Salmonella* and *Shigella* | ◯ | ◯ | ◯ | ◯ |

Q8. If there is an organism you consider high priority that wasn't listed in Q7, please list it here

[ ]

Q9. Phage therapy should be available that is selective, individualised and targets specific microorganisms (bespoke phage therapy). *

1 2 3 4 5

Strongly disagree Strongly agree

Q10. Phage therapy should be readily available that possesses a broad spectrum of action, targeting multiple microorganisms likely to be responsible for a clinical syndrome or condition (phage cocktail). *

1 2 3 4 5

Strongly disagree Strongly agree

Q11. In your opinion, how would you rank the following clinical syndromes in terms of their priority for phage therapy? *

| | High priority | Medium priority | Low priority | Uncertain |
|---|---|---|---|---|
| Bone and joint infection including prosthetic device related infection | ○ | ○ | ○ | ○ |
| Cystic fibrosis | ○ | ○ | ○ | ○ |
| Diabetic foot infection | ○ | ○ | ○ | ○ |
| Gastrointestinal tract infection | ○ | ○ | ○ | ○ |
| Infection in burns patients | ○ | ○ | ○ | ○ |
| Infections in transplant and immunosuppressed patients | ○ | ○ | ○ | ○ |
| Infective endocarditis and cardiac device infections | ○ | ○ | ○ | ○ |
| Intra-abdominal infection including intra abdominal abscesses | ○ | ○ | ○ | ○ |
| Pulmonary infection (excluding CF) | ○ | ○ | ○ | ○ |
| Sepsis/bacteremia | ○ | ○ | ○ | ○ |
| Skin and soft tissue infection | ○ | ○ | ○ | ○ |
| Urinary tract infections | ○ | ○ | ○ | ○ |

Question 11a. If there are any clinical syndromes not listed above that you feel are a high priority, please list them here.

Q12. In the past year, have you come across a patient who you think may have benefited from phage therapy? *

- ◯ Yes, multiple patients
- ◯ Yes, a single patient
- ◯ No
- ◯ I am unsure

Q13. Which administration routes would you find useful for administering phage therapy? (Please select all that apply) *

- ☐ Inhalation
- ☐ Instillation (e.g., into organs or cavity)
- ☐ Intravenous
- ☐ Oral
- ☐ Phage-coated devices (including prostheses and indwelling catheters)
- ☐ Topically
- ☐ Other

Q13a. As you selected other, please specify which other administration routes you would find useful for administering phage therapy.

____________________

Q14. Do you think phage therapy delivery protocols should include therapeutic phage monitoring? (e.g., Similar to therapeutic drug monitoring) *

(1) (2) (3) (4) (5)

Strongly disagree — Strongly agree

Q15. Do you think Australia's national Antimicrobial Resistance (AMR) action plan is adequate to control the spread of AMR in Australia? *

- ◯ Yes
- ◯ No

Q16. Do you believe phage therapy is likely to be an effective solution to AMR? *

- ◯ Yes
- ◯ No

| | |
|---|---|
| **From:** | |
| **To:** | |
| **Cc:** | Stephanie Lynch |
| **Subject:** | 2022/PID00489 - 2022/ETH00432: Application HREA - Approved |
| **Date:** | Wednesday, May 18, 2022 3:00:13 PM |

Date of Decision Notification: 18 May 2022

Dear Professor Jonathan Iredell,

Thank you for submitting the following Human Research Ethics Application (HREA) for HREC review;

2022/PID004892022/ETH00432: Perception of phage therapy among Australian authorised prescribers

Thank you for your correspondence addressing the matters raised in the HREC's letter following single ethical review of the above project at its meeting held on 5 April 2022. The project was determined to meet the requirements of the National Statement on Ethical Conduct in Human Research (2007) and was APPROVED.

This email constitutes ethical and scientific approval only. This project cannot proceed at any site until separate research governance authorisation has been obtained from the Institution at which the research will take place.

This project has been Approved to be conducted at the following sites:

- Westmead Hospital
- Westmead Institute for Medical research

The following documentation was reviewed and is included in this approval:

1. HREA Application
2. Protocol, version 1 dated 7 March 2022
3. Participant Information and Consent Form, version 2 dated 20 April 2022
4. Perception of phage therapy among Australian authorised prescribers
5. Perception of phage therapy data collection form
6. Prescriber survey distribution email
7. Prescriber survey questionnaire

Application Documents - (link will only be active for 14 days from the decision date. The approved documents are also available to download from forms section of this project in REGIS)

The approval is for a period of 5 years from the date of this e-mail (18 May 2022)

The Coordinating Principal Investigator will:
• provide the HREC with an annual report and the final report when the project is

completed at all sites. This will be through the submission of a milestone in REGIS.
• immediately report anything that might warrant review of ethical approval of the project.
• submit proposed amendments to the research protocol, including; the general conduct of the research, changes to CPI or site PI, an extension to HREC approval, or the addition of sites to the HREC before those changes can take effect. This will be through a notification of an amendment in REGIS
• will notify the HREC if the project is discontinued at a participating site before the expected completion date, with reasons provided.

Submission of annual progress/final reports (milestone), amendments and safety reports should be done through the forms provided in REGIS. Guidance on these processes can be found on the REGIS website.

It is noted that the Western Sydney Local Health District Human Research Ethics Committee is constituted in accordance with the National Statement on Human Conduct in Human Research, 2007 (NHMRC).

Please contact us if you would like to discuss any aspects of this process further, as per the contact details below. We look forward to managing this study with you throughout the project lifecycle.

Regards,
WSLHD Research Office
